# Evaluating Genome Sequencing Strategies: Trio, Singleton, and Standard Testing in Rare Disease Diagnosis

**DOI:** 10.1101/2024.12.20.24319228

**Authors:** Daniel Kaschta, Christina Post, Franziska Gaass, Bianca Greiten, Anna-Sophie Liegmann, Rebecca Gembicki, Jelena Pozojevic, Michelle Meyenborg, Janne Wehnert, Katharina Schau-Römer, Franka Rust, Maj-Britt Salewski, Kristin Schulz, Varun Sreenivasan, Saranya Balachandran, Kristian Händler, Veronica Yumiceba, Laelia Rösler, Andreas Dalski, Kirstin Hoff, Nadine Hornig, Juliane Köhler, Vincent Arriens, Caroline Utermann-Thüsing, Kimberly Roberts, Eva Maria Murga Penas, Christine Zühlke, Monika Kautza-Lucht, Maike Dittmar, Irina Hüning, Yorck Hellenbroich, Britta Hanker, Valerie Berge, Friederike Birgel, Philip Rosenstiel, Andre Franke, Janina Fuß, Britt-Sabina Löscher, Sören Franzenburg, Dzhoy Papingi, Amelie van der Ven, Sandra Wilson, Rixa Woitschach, Jasmin Lisfeld, Alexander E. Volk, Theresia Herget, Christian Schlein, Anna Möllring, Birga Hoffmann, Imke Poggenburg, Henning Nommels, Milad Al-Tawil, Gloria Herrmann, Andreas Recke, Louiza Toutouna, Olaf Hiort, Nils Margraf, Bettina Gehring, Hiltrud Muhle, Tobias Bäumer, Lana Harder, Alexander Münchau, Norbert Brüggemann, Inga Vater, Almuth Caliebe, Inga Nagel, Malte Spielmann

## Abstract

**Purpose:** Short-read genome sequencing (GS) is a comprehensive genetic testing method capable of detecting multiple variant types. Despite its technical advantages, systemic comparisons of singleton GS (sGS), trio GS (tGS), and exome sequencing-based standard-of-care (SoC) in real-world diagnostics remain limited.

**Methods:** We systematically compared sGS, tGS, and SoC genetic testing in 448 patients with rare diseases in a blinded, prospective study. Three independent teams evaluated the diagnostic yield, variant detection capabilities, and clinical feasibility of GS as a first-tier test. Diagnostic yield was assessed through both prospective and retrospective analyses.

**Results:** In prospective analyses, tGS achieved the highest diagnostic yield for likely pathogenic/pathogenic variants (36.8%) in a newly trained team, outperforming the experienced SoC team (36.0%) and the sGS team (30.4%). Retrospective analyses, accounting for technical variant detection and team experience differences, reported diagnostic yields of 38.6% for SoC, 41.3% for sGS, and 42.2% for tGS. GS excelled in identifying deep intronic, non-coding, and small copy-number variants missed by SoC. Notably, tGS additionally identified three de novo variants classified as likely pathogenic based on recent GeneMatcher collaborations and newly published gene-disease association studies.

**Conclusion:** GS, particularly tGS, demonstrated superior diagnostic performance, supporting its use as a first-tier genetic test. sGS offers a cost-effective alternative, enabling faster, more efficient diagnoses for rare disease patients.

## Introduction

Advances in healthcare have steadily contributed to a global decline in disease burden.^1^ With growing insights into the mechanism and etiology of diseases, recent studies reveal that genetic factors outweigh environmental contributions in approximately 40% of disease phenotypes.^2^ Notably, in cases of infant mortality, genetic associations have been identified in 41% of instances, primarily linked to rare genetic variants^3^. Rare diseases, in general, represent a significant public health burden due to their complexity, limited treatment options, and high diagnostic and therapeutic costs.^4^ In Germany, approximately four million people are affected.^5^ In Western populations, between 6,000 and 10,000 distinct rare diseases impact 3.5–5.9% of individuals, with over 80% believed to have a genetic origin.^6,7^ The combination of traditional diagnostic methods, including karyotyping, array comparative genomic hybridization (array-CGH), and exome sequencing (ES), remains the standard of care (SoC) for establishing a broad range of molecular diagnoses. Although effective, when used together these methods are time-consuming and still face significant limitations in detecting non-coding variants, smaller structural variants, and complex genomic rearrangements due to inherent limitations in resolution.^8,9–11^ Short-read genome sequencing (GS) presents a promising alternative to these traditional methods, offering a more comprehensive strategy.^12^ Unlike targeted approaches, GS provides a more complete genomic view, enabling the improved detection of a broader spectrum of variants, including those in non-coding regions, intronic variants, short tandem repeats (STR), and copy-number variants (CNV).^13–15^ Recent publications show overall retrospective detection rates of up to about 95% for the majority of known clinical indications.^16,17^ This makes GS a potentially very powerful unifying tool that could enhance diagnostic yield and reduce the time to diagnosis.^18^ A recent review including 27 studies places the diagnostic yield of GS as a first-line test in rare disease at about 45%.^19^ However, larger prospective studies focusing on the benefits of GS in a real-world setting are still scarce and do not evaluate all potential frameworks.^19,20^ In this clinically heterogeneous study we aim to fill this gap by directly comparing the diagnostic yields of singleton genome sequencing (sGS), trio genome sequencing (tGS), and SoC to verify whether short read GS can truly serve as a “one-test-for-all” strategy for rare disease cases. We focus on GS effectiveness as comprehensive diagnostic tools, capable of identifying a wide variety of genomic variants potentially missed by conventional SoC methods. Furthermore, our prospective and retrospective results highlight the feasibility of the three methods within the real-world setting of the second-largest university hospital in Germany.^21^ We conducted a comparative analysis focusing on diagnostic yield, implementation processes, and overall practicality to determine which approach offers the optimal balance of diagnostic comprehensiveness and economic sustainability.^22^

## Methods

### Cohort and Study Subjects

Between January 2022 and April 2023, the Universitätsklinikum Schleswig-Holstein (UKSH) Genome Consortium recruited 448 patients with rare diseases including developmental disorders, congenital abnormalities, syndromic conditions, or rare neurological syndromes, and applied short-read GS alongside SoC diagnostic strategies. Other inclusion criteria were a high likelihood of a genetic etiology as determined by a clinical geneticist, no pre-existing clinical genetic diagnosis, no prior SoC or GS, eligibility to receive SoC covered by insurance, samples meeting technical quality control standards, and provision of full consent to participate in the study. Each index patient categorized with a syndromic or developmental disorder underwent SoC testing, including sequential analyses using karyotyping, array-CGH, and ES. If karyotyping or array-CGH identified a causal variant, ES was not performed. For other disease categories, such as neurological disorders, SoC included at least ES. Additionally, GS was conducted on index patients and available family members, including parents and siblings. In the following, we will label all included singleton cases as singletons (84 cases) while all multi-family member cases, regardless of size, will be grouped under trios (332 cases).

### Analysis Strategies

In this prospective, blind study three independent analysis teams analyzed the cases:

● The SoC team analyzed cases based on the phenotypic indications of the index patients’ karyotype, array-CGH, and ES data.
● The sGS team analyzed cases based on the phenotypic indications and the index patients’ GS data.
● The tGS team focused only on trio cases and analyzed them based on the phenotypic indications, the GS data of the index patients, and available siblings or parental samples.

Following their unique approach, each team committed to logging in only their candidate results without knowing the results of the other groups. The results were collected and assessed by a collaborative evaluation committee comprising representatives from each team, who determined the final diagnostic outcome reported to the patient.

Our three strategies were compared across two stages. The first stage, termed the prospective stage, involved evaluating prospective data submitted by each team. The prospective diagnostic yield refers to each team’s ability to identify the variants ultimately confirmed as the final diagnostic result by the committee.

The second stage, referred to as the retrospective stage, involved all teams evaluating the final results retrospectively. This stage aimed to determine whether the identified variants were technically recognized and, if so, whether they passed the analysis filters used in the different approaches (SoC, sGS, and tGS). The retrospective diagnostic yield reflects the capability of each diagnostic strategy to identify the variants, independent of team-specific experience.

### Genome Sequencing and Sample Quality Control

A total of 1,148 DNA samples derived from patient blood underwent standard library preparation protocols using the DNA PCR-Free Prep, Tagmentation-Kit (Illumina, Inc., San Diego, CA, USA). This was followed by short-read GS with 16 samples per NovaSeq Flow Cell (Illumina, Inc.) on a NovaSeq™ 6000 instrument (Illumina, Inc) using the NovaSeq 6000 S4 Reagent Kit (Illumina, Inc) with 2 x 150 bp paired-end reads and an anticipated genome-wide coverage (mean mapped read depth) of 29-fold minimal; actual mean coverage of 38 × was documented. Due to technical issues or low sample quality, sequencing was repeated for 68 samples (6.55%). Adapting to a higher DNA concentration, transitioning from an initial 300 ng to 600 ng, substantially improved assay robustness.

### Exome Sequencing and Sample Quality Control

For ES the coding regions of 19,433 genes were enriched using the Nextera Flex for Enrichment kit (Illumina, Inc.) and the xGen Exome Research Panel v2 (Integrated DNA Technologies, Inc.Coralville, IA, USA). Sequencing of the resulting libraries was conducted on the NovaSeq™ 6000 platform with 2 x 150 bp paired-end reads. Raw data were aligned to the human reference genome GRCh38 (hg38). The mean sequencing depth for index/mother/father samples was 202.8×/159.1×/155.2×, with 97.8%/97.0%/97.2% of target regions covered at least 20 × and 97.4%/96.3%/96.5% covered at least 30 ×.

### Genome and Exome Variant Interpretation

Variant detection for genomes was performed using the Germline Pipeline of the DRAGEN™ (Dynamic Read Analysis for GENomics) Bio IT platform v3.7.5 (Illumina, Inc.), Manta Structural Variant Caller (Illumina, Inc.) for CNV detection and ExpansionHunter(Illumina, Inc.) for detection of STRs. ES analysis was performed using the IKMB DRAGEN Pipeline (https://github.com/ikmb/dragen-variant-calling), based on the commercial DRAGEN Pipeline v3.10.4 (Illumina, Inc.). Variants with coverage below 20 × or with less than 20% variant reads were excluded.

GS data was interpreted using the TruSight Software Suite v2.6 (TSS, Illumina, Inc.) while for ES data, variant analysis was performed using Alissa Interpret v5.4.2 (Agilent Technologies, Santa Clara, CA, USA). Variant calling was followed by the application of a series of filtering steps for variant prioritization. For rare diseases, variants with allele frequencies below 1% for recessive inheritance and 0.1% for dominant inheritance based on the frequencies from the Genome Aggregation Database (gnomAD)^25^ and the Exome Aggregation Consortium (ExAC)^26^ were considered as well as known disease-causing variants from the literature.

Protein-altering variants and those affecting canonical splice sites (for ES: +/-10 bp) were assessed for functional relevance, conservation, and splicing impact using various *in silico* tools (e.g., CADD score). For genome-wide strategies, variants were evaluated across the entire genome. All candidate variants were reported if alignment matched with the analyzed individual’s symptoms and inheritance pattern. Additionally, the tGS team tracked potentially promising *de novo* variants with unknown gene-disease mechanisms as scientific findings, prioritizing them for future re-evaluation or submission to GeneMatcher databases. All variants were associated with the patient’s phenotype using Human Phenotype Ontology (HPO) terms and evaluated using databases such as OMIM, PubMed, ClinVar, Decipher, and *in silico* prediction tools (e.g., CADD score).

Allele frequencies were assessed using the Genome Aggregation Database (gnomAD v3.1.2). Visual validation of variants was performed using the Integrated Genome Viewer (IGV; Version: 2.13.2-2.17.4).

### Genome and Exome Variant Classification

Variant classification followed the guidelines of the American College of Medical Genetics and Genomics (ACMG) and Association for Molecular Pathology (AMP) and the Association for Clinical Genomic Science (ACGS).^23,24^ The diagnostic yield is categorized into three classifications:

1. **P/LP**: Pathogenic or likely pathogenic variants were identified as the main clinical indication that align with the patient’s phenotype and inheritance pattern.
2. **VUS**: Variants of uncertain significance were assessed as the main clinical indication that matched the patient’s phenotype and inheritance pattern, supported by a high level of evidence (“Hot VUS”).
3. **No Findings**: No diagnostic variants were identified.

Moreover, the genomic data of the sequenced individuals (patients and relatives) was analyzed to report for P/LP variants classified as secondary findings (SF) within actionable genes curated in the ACMG SF list v3.2.^27^

### Array-CGH and Sample Quality Control

DNA was isolated from patient blood samples, with approximately 1µg of DNA per sample. These samples were subjected to array-CGH using the Agilent SurePrint G3 Human CGH Microarray Kit (8×60K) (Agilent, Inc.). Standard protocols for DNA labeling and hybridization were followed, with scans performed on the Agilent DNA Microarray Scanner.

### Array-CGH Data Analysis

Array CGH data was processed using Genomic Workbench software (v7.0) (Agilent, Inc.), which facilitated the detection of CNVs. CNV interpretation was performed by comparing identified variants to known genomic databases, including ClinGen, DECIPHER, and OMIM, with additional *in silico* analysis performed to assess potential pathogenicity. The results were cross-referenced with patients’ clinical phenotypes to determine relevance and potential clinical significance (Variants classified as P/LP and strong VUS combined).

### Karyotype Analysis

Metaphases of each case were G-banded with conventional trypsin-Giemsa staining and analyzed at 400 or 550 band level according to referral reason.^28^ Karyotypes were described following the guidelines of the International System for Human Cytogenetic Nomenclature (ISCN 2020).^29^ Digital image acquisition, processing, and evaluation were performed using NEON interface for case and image data software version 1.3 (MetaSystems, Altussheim, Germany). Chromosomal abnormalities, such as aneuploidies, translocations, deletions, and duplications, were identified and classified. Findings were interpreted in the context of clinical phenotypes, with cross-referencing against established cytogenetic databases (e.g., DECIPHER, OMIM) to assess clinical significance.

### Comparison and Learning Curve

Case comparisons between teams began after the first 100 cases, with results subsequently shared among the groups. This approach enabled continuous error identification and inferred adjustments to minimize disparities stemming from potential experience gaps within the teams. To illustrate this process, the prospective diagnostic yield was tracked using a sliding window analysis, capturing temporal changes in performance and optimization. A window size of 25 cases before and after each case was selected to calculate the average diagnostic yield for each team based on the total number of cases analyzed. These learning and adaptation curves, displayed with standard deviation, were used to monitor trends and evaluate the performance of each strategy across all 332 trio cases.

## Results

### Diagnostic Yield

Between January 2022 and April 2023, 1,148 patients and their relatives were recruited and sequenced, resulting in 416 index cases (332 trios and 84 singletons). Of the sequenced individuals, 1,006 genomes met inclusion criteria, while 32 cases were excluded. The cohort included 84 singletons, 50 duos, 261 trios, nine quartets, a quintet, and a sextet. Seven quartet families and the sextet family involved two affected patients, while the quintet family involved three. Males comprised 66.3% of the cohort, and 84.9% of patients were under 20 years of age. Patients were categorized by disease groups to explore potential distinctions(**Figure 1**). The initial evaluation of the 332 trio cases revealed the following diagnostic yields for P/LP variants: 36.0% (n = 119 cases) for SoC, 30.4% (n = 101 cases) for sGS, and 36.8% (n = 122 cases) for tGS. When including VUS, the combined percentage of cases with clinically significant variants increased to 46.5% (n = 159 cases) for SoC, 35.2% (n = 117 cases) for sGS, and 42.6% (n = 141 cases) for tGS. For the evaluation of 84 singleton cases, the differences between SoC and sGS were smaller. SoC identified P/LP variants in 39.2% of cases (n = 33) and VUS in 21.4% of cases (n = 18), yielding a total diagnostic rate of 60.7%. In comparison, sGS detected P/LP variants in 36.9% of cases (n = 31) and VUS in 10.1% (n = 8 cases), for a total diagnostic yield of 47.0% (**Figure 2**).

**Figure 1.**
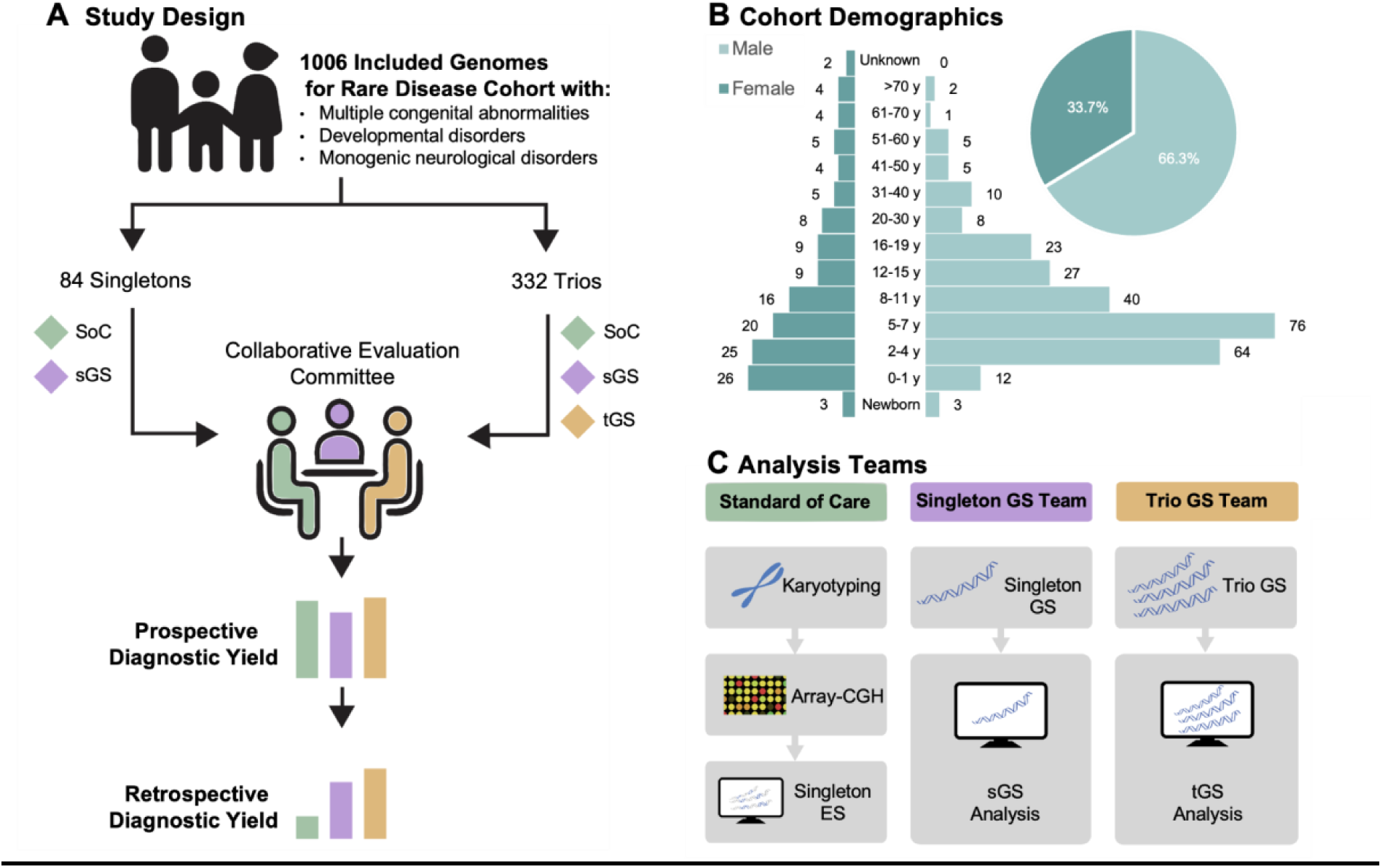
Study Design and Cohort Overview for Rare Disease Analysis. **(A)** In this study our cohort comprised 1,006 individuals from 416 index family cases of rare diseases. Among these, 84 singletons were analyzed by the Standard of Care (SoC) and singleton genome sequencing (sGS) team, while 332 trio cases (multi-family member cases) received additional GS for family members and were further analyzed by the trio genome sequencing (tGS) team. A collaborative evaluation committee assessed both prospective and retrospective results. **(B)** The demographic overview illustrates the study sex and age distribution of the 276 male and 140 female patients. **(C)** Three analysis strategies were employed: SoC (sequential testing of index blood samples with karyotyping, array-CGH, and exome sequencing), sGS (genome sequencing of the index patient), and tGS (genome sequencing of the index patient with parental and sibling data when available).

**Figure 2.**
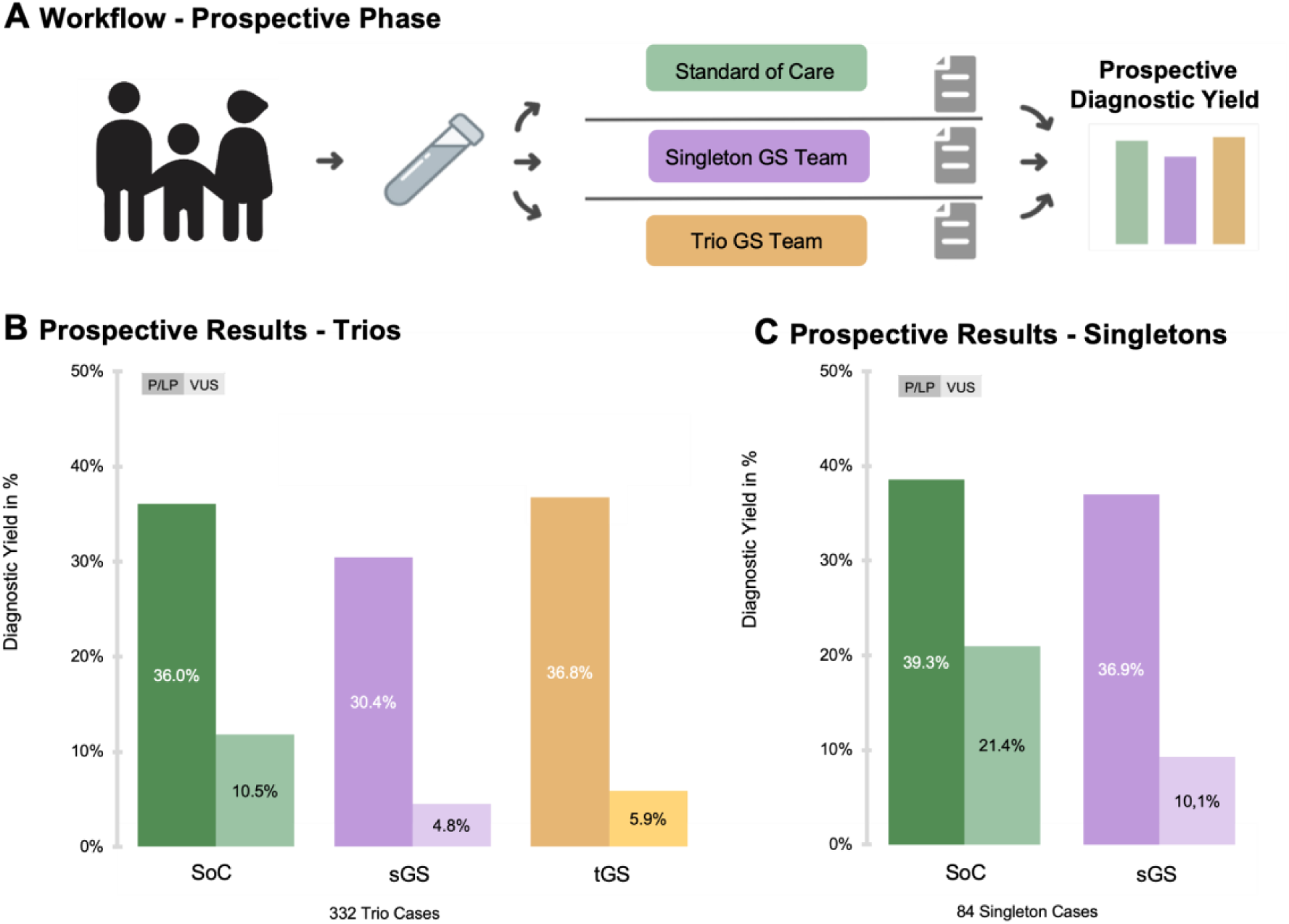
Prospective Workflow and Diagnostic Yield Across Three Analysis Strategies. **(A)** Workflow for the analysis of prospective data with evaluation by the collaborative evaluation committee. **(B)** Prospective diagnostic yield for 332 trio cases, comparing the standard of care (SoC), singleton genome sequencing (sGS), and trio genome sequencing (tGS) strategies, with a focus on pathogenic or likely pathogenic (P/LP) variants and variants of uncertain significance (VUS). **(C)** Prospective diagnostic yield for 84 singleton cases, comparing the SoC and sGS strategies, focusing on P/LP variants and VUS.

The prospective evaluation stage was followed by a retrospective assessment of results to address experience gaps in evaluating the strategies. The curated data for all 332 trio cases revealed a retrospective diagnostic yield for P/LP variants of 38.6% (n = 128 cases) for SoC, 41.3% (n = 137 cases) for sGS, and 42.2% (n = 140 cases) for tGS. The percentage of VUS was consistent across all strategies at 10.8% (n = 17 cases). Including the VUS category, the combined percentage of cases with a variant with clinical significance was 49.6% (n = 145 cases) for SoC, 52.3% (n = 154 cases) for sGS, and 53.2% (n = 157 cases)for tGS. In the retrospective analysis of the 84 singleton cases, SoC and sGS demonstrated identical performance, with 39.5% (n = 32) for P/LP variants and 21% (n = 17 cases) for VUS. For these cases, no variants were exclusive to either approach (**Figure 3**).

**Figure 3.**
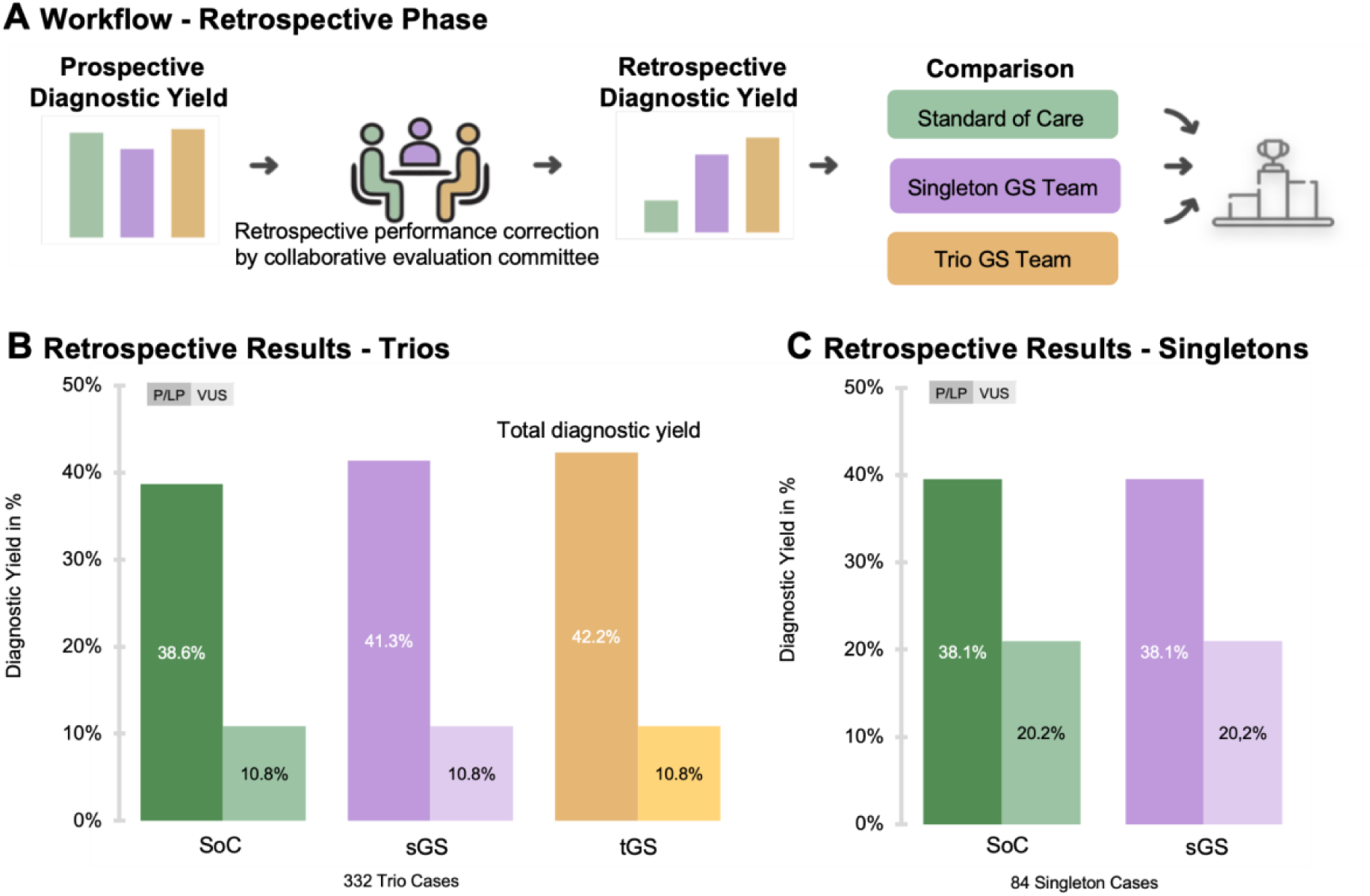
Retrospective Diagnostic Yield. **(A)** Workflow for the retrospective analysis of prospective data following evaluation by the collaborative evaluation committee. **(B)** Retrospective diagnostic yield for 332 trio cases, comparing the standard of care (SoC), singleton genome sequencing (sGS), and trio genome sequencing (tGS) strategies, with a focus on pathogenic or likely pathogenic (P/LP) variants and variants of uncertain significance (VUS). The tGS covered all identified variants (total diagnostic yield) **(C)** Retrospective diagnostic yield for 84 singleton cases, comparing the SoC and sGS strategies, focusing on P/LP variants and VUS.

### GS Unique Variants

A total of 229 cases, singletons and trios combined, were identified with a diagnostic genetic cause (P/LP and VUS) based on the final retrospective results. According to ACMG/AMP criteria, 173 cases had variants classified as P/LP, while 56 cases were classified with VUS. In total, 191 disease-causing variants were identified **(Table S1 in the Supplementary Appendix)**, 180 of which were detectable using the SoC approach, with 12 additional variants exclusively identified through the GS pipeline (**Figure 4A**).

**Figure 4.**
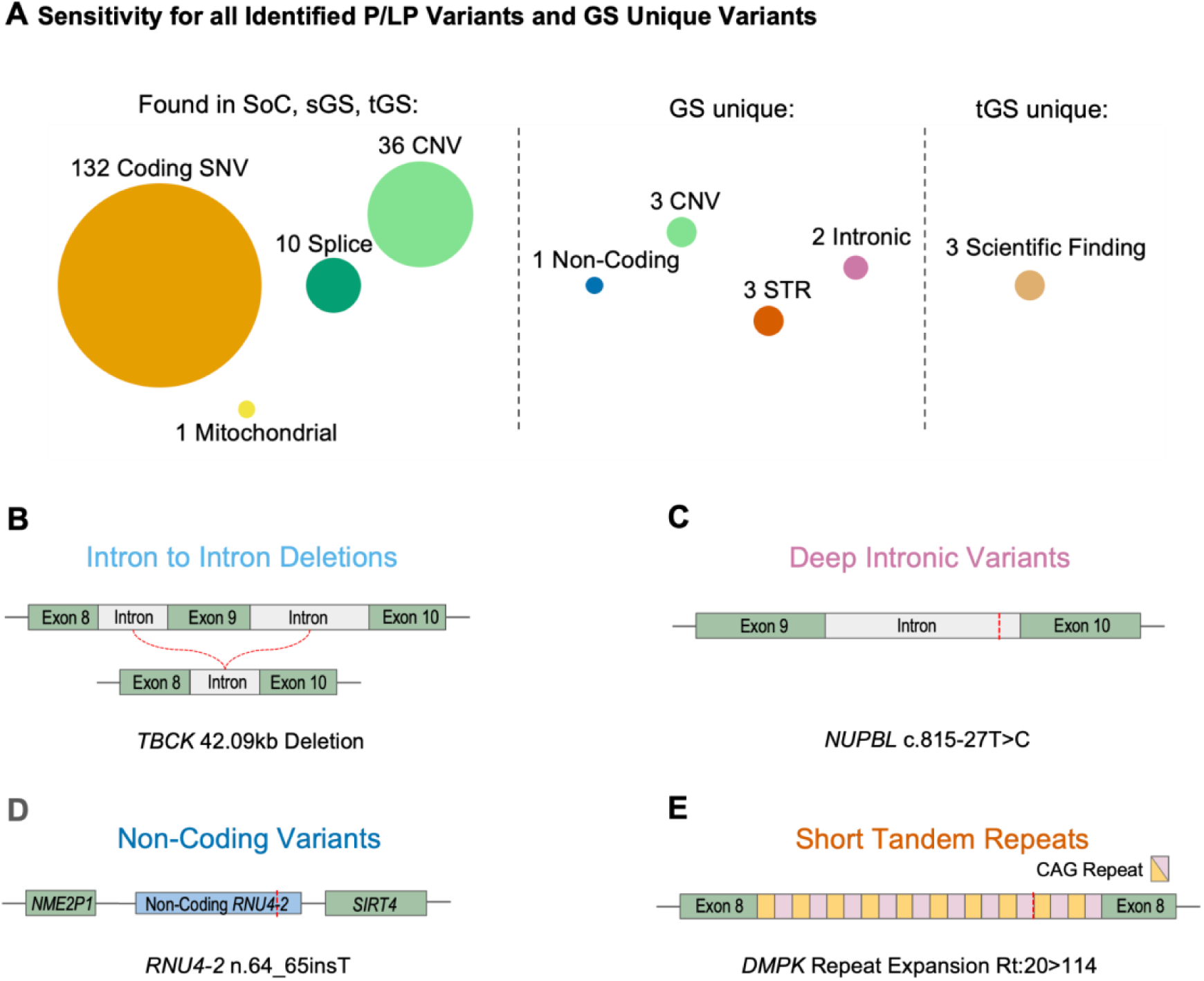
Strategy Performance for Variant Types and Unique Genome Variants. **(A)** The sensitivity for detecting specific variant types varied across the tested strategies. Of the 191 pathogenic or likely pathogenic (P/LP) variants identified within the cohort, 180 were detected using the standard of care (SoC), singleton genome sequencing (sGS), and trio genome sequencing (tGS) strategies, encompassing single-nucleotide variants (SNVs), copy-number variants (CNVs), splice variants, and mitochondrial variants. Unique variant types identified throughGS included additional intron-to-intron CNVs, deep intronic variants, non-coding variants, and short tandem repeats (STRs). Notably, tGS identified three additional *de novo* variants, later reclassified as P/LP based on GeneMatcher collaborations and emerging studies linking new gene-disease associations. **(B-E)** Representative examples include an intron-to-intron deletion (CNV), a deep intronic variant, a non-coding variant, and an STR repeat tract expansion (Rt) detected using GS strategies.

The variants covered by all strategies included 36 CNVs, 133 coding SNVs, 10 splice variants, and one mitochondrial variant. Among the GS unique variants, we detected nine variants in total by sGS and tGS: three intragenic CNVs spanning from one intron to another intron, three STRs, two intronic variants, and one non-coding variant. Unique to the tGS strategy we discovered three *de novo* variants that initially lacked a gene-disease association but could later be upgraded to LP via a GeneMatcher cooperation with similar patients.^30^ GS identified STRs in three patients: two with estimated CAG repeat expansions of 143 and 114 in the *DMPK* gene, and one with a CGG repeat expansion of 90 in the *FMR1* gene. (**Figure 4B-E**). During the study, publications on recently discovered pathogenic variants within the non-coding snRNA *RNU4-2* locus highlighted the potential of GS strategies for reanalysis.^31^ We focused on the recently uncovered disease potential of snRNAs from the *RNU* family. A comprehensive VCF reanalysis of the full cohort was conducted, targeting specifically genomic regions for *RNU4-2*, *RNU2-2*P, *RNU5A-1*, and *RNU5B-*1. This approach enabled the mapping and classification of potential variants. Notably, we identified a patient carrying the most common previously described disease-causing variant in the *RNU4-2* critical region, n.64-65insT, which is associated with impaired RNA splicing and contributes to the patient’s neurological symptoms **(Supplementary Appendix Figure S2)**.^32^

### Patterns of Inheritance and Variant Types

From the 118 trio cases in which we identified P/LP variants responsible for the individuals’ phenotypes and both parental genomes were available, we assessed 114 unique variants and their patterns of disease inheritance. Here, 64% of the inheritance patterns were *de novo*, 15.8% maternally or paternally inherited heterozygous, 9.6% were biparental compound heterozygous, 8.8% biparental homozygous and 1,8% maternal hemizygous (**Figure 5A**).

**Figure 5.**
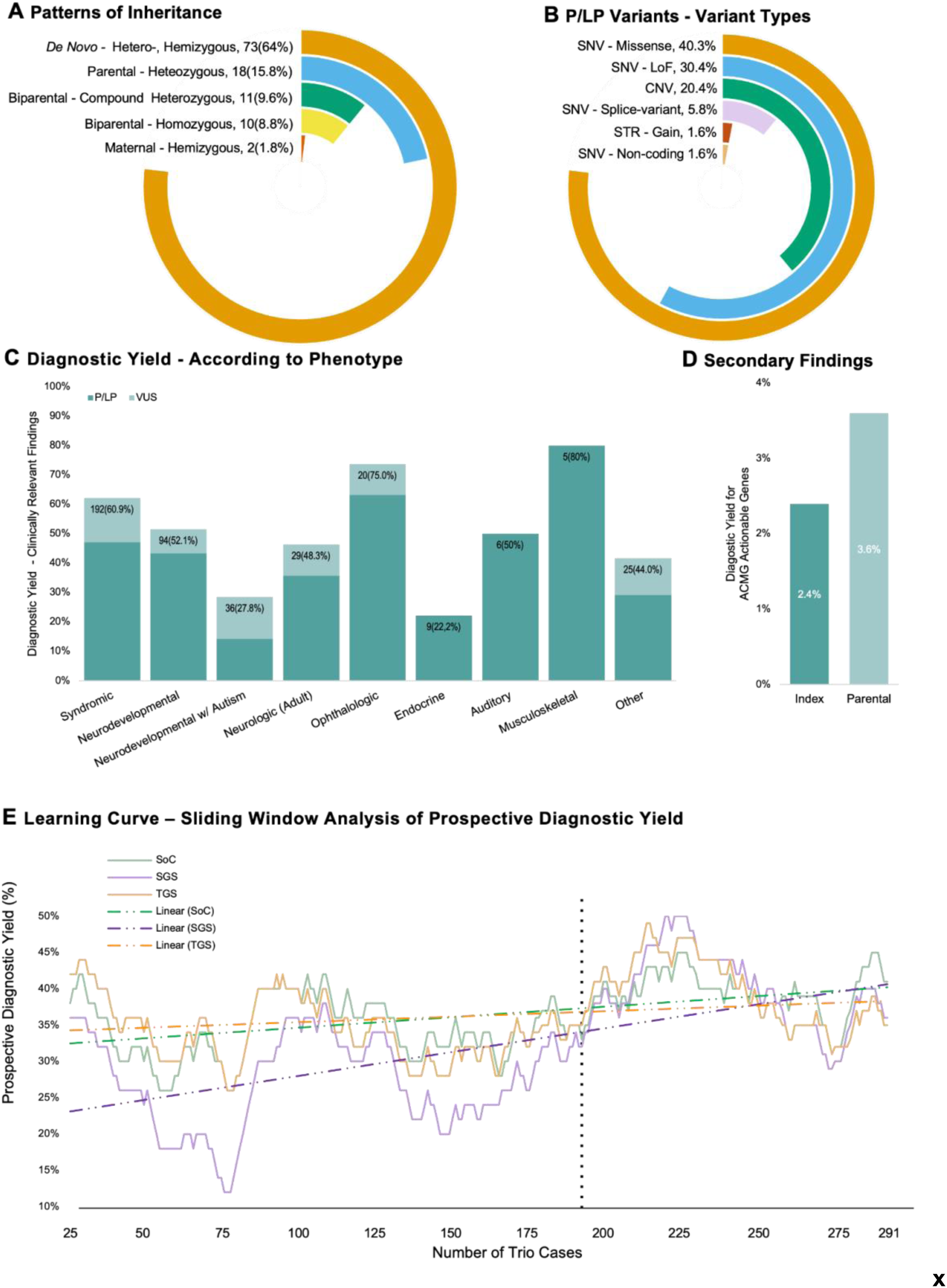
Inheritance and Variant Types / Disease Categories / Learning Curve. **(A)**The distribution of inheritance patterns for genetically identified diseases solved with pathogenic or likely pathogenic (P/LP) variants. **(B)** All 191 P/LP variants are classified by type, including missense small nucleotide variants(SNVs), loss-of-function (LoF) SNVs, splice-effect SNVs, non-coding SNVs, short tandem repeats (STRs), and copy-number variants (CNVs). **(C)** Retrospective diagnostic yield is presented for various disease categories within the cohort, categorized based on the final diagnostic outcome. **(D)** The percentage of trio cases with P/LP variants classified as secondary findings (SF) based on the ACMG SF 3.2 list, encompassing findings in either the index patient or analyzed parents. **(E)** A sliding window analysis (25 cases before and after each case) evaluates the prospective diagnostic yield for trio cases with P/LP variants. The comparison includes standard of care (SoC), singleton genome sequencing (sGS), and trio genome sequencing (tGS), with a dotted line at 190 cases representing a symbolic break-even point for all strategies, defined as a 1% standard deviation difference.

Among the 191 P/LP variants identified, single-nucleotide variants (SNVs) were the most common, comprising 40.3% missense variants, 30.4% loss-of-function (stop gain, start loss, frameshift), 20.4% CNVs, 5.8% splice-region SNVs, and 1.6% intronic or non-coding SNVs. STRs also represented 1.6% (**Figure 5B**).

### Disease Categories and Their Respective Diagnostic Yields

Syndromic (n = 192), neurodevelopmental (n = 130), and neurological (n = 29) cases represented the largest proportion within our cohort. The retrospective diagnostic yield for clinically significant findings in syndromic cases was 60.9%. However, this yield varied significantly across different disease categories, highlighting differences in diagnostic outcomes. This was particularly evident in the 36 cases of neurodevelopmental disorders involving autism, which had a combined diagnostic yield of 27.8% for clinically significant variants, compared to the 94 cases of neurodevelopmental disorders without autism, which demonstrated a yield of 52.1% (**Figure 5C**).

### Secondary Findings

In our cohort, SF in genes included in the ACMG SF v3.2 list for reporting SF in clinical exome and genome sequencing were identified in 2.4% of 332 trio cases.^27^ In contrast, the prevalence of identified SF in parents was only observed at 3.6% (**Figure 5D**).

### Prospective Implementation of GS in the Clinic

Of the three independent teams that analyzed the cases in this study, the SoC team comprised experienced members from the diagnostic department, highly proficient in routine analytical workflows such as karyotyping, array-CGH, and ES. The GS teams included analysts with only basic training in GS analysis, leading to comparatively less familiarity with the GS pipeline. Additionally, the GS pipeline itself was newly introduced and lacked the extensive validation and optimization that the SoC pipeline had undergone.

During the study, the tGS team reported 255 candidate variants across 332 trio cases, compared to 312 reported by the SoC team and 414 by the sGS team. In the 84 singleton cases, 69 candidate variants were identified by the SoC team, while 110 were reported by the sGS team, highlighting variability in candidate reporting between teams with differing levels of expertise, access to analytic data supporting variant identification, and adaptation to analytical methods.

To address heterogeneity in team experience and monitor the optimization process, the diagnostic yield of the three approaches was continuously evaluated and openly reviewed by a collective evaluation committee beginning after the first 100 cases. A sliding window visualization revealed that after 76 cases, the diagnostic yield was 12% for the sGS team, compared to 36% for the tGS team and 34% for the SoC team. Despite this initial disparity, the sGS team exhibited the steepest improvement, eventually matching the performance of the other teams after 190 cases (within a 1% standard deviation) and briefly surpassing them after 220 cases, likely due to the steadily increasing experience and enhanced workflow efficiency, reflecting a successful adaptation to the initial gaps in experience and pipeline integration. The tGS approach, enhanced by the inclusion of additional inheritance data for candidate variants, demonstrated the highest yield closely followed by the SoC framework (**Figure 5E**).

## Discussion

In this study, we compared short-read GS (sGS and tGS) to SoC methods for diagnosing rare diseases. The tGS team had the highest diagnostic yield with P/LP variants in 42.3% of cases, followed by sGS at 41.4%, and both outperforming SoC at 38.7%. GS’s broader coverage and inheritance data improved the identification of complex structural and non-coding variants. Additionally, tGS uncovered *de novo* variants that were initially considered candidate causal variants and were subsequently validated as disease-causing through GeneMatcher collaborations or literature review. These results align with Tan et al.’s comparison of singleton and trio ES, which demonstrated improved detection of causal *de novo* variants without established gene-disease associations. Notably, their study involved a relatively small cohort of 30 rare disease cases, demonstrating a 3.3% increase in diagnostic yield with the trio approach.^33^

Previous studies have suggested that GS has the potential to serve as a universal diagnostic tool for rare diseases. Our data confirm this for tGS in this prospective real-world study, aligning well with the findings of Schobers et al. that GS effectively functions as a “one-test-for-all” strategy.^16^ This referenced study reported that GS retrospectively detected 94.9% of 1,271 clinically relevant variants in 1,000 cases, covering small, large, and structural variant types.

On the other hand, sGS, though slightly lower in performance than tGS, suggests that including parental data to interpret *de novo* variants resulted only in a minor diagnostic benefit, particularly when considering the substantial additional costs associated. This makes sGS a cost-effective alternative to tGS with a high diagnostic yield. However, short-read GS approaches face limitations, particularly in mapping complex genomic regions such as repetitive sequences, segmental duplications, or specifically long STRs.^16^ Here, GS demonstrated potential as a nonspecific primary screening method by indicating a possible increase in motif length of a specific repeat region. However, additional diagnostic approaches, such as long-read GS, are required to accurately determine the exact motif length and therefore provide a more comprehensive understanding of the genomic variation.

One positive by-product of tGS is its ability to identify SF in ACMG-designated actionable genes in both the index patient and the parents. These findings can guide preventive care and early interventions, such as cancer screenings or lipid-lowering therapies, thereby reducing morbidity and mortality. In this study, 2.4% of index patients and 3.6% of trio-case parents had actionable SF. For the parents this is slightly lower than anticipated based on prior studies.^34^ This discrepancy can be attributed to incomplete parental data in 50 duo cases out of the 332 trio cases.

An increasing challenge in cases with inconclusive findings is the growing demand for the reanalysis of genomic data. In particular, automated processes have been proposed as a means to streamline and enhance the reanalysis of patient genomic data.^35^ A recent meta-analysis of 29 studies has shown that reanalyzing ES and GS data starting approximately two years or later after the initial analysis can provide conclusive findings for about 10% of previously inconclusive cases.^36^ This study underscores the key advantage of GS as a potential alternative to SoC, partly because of its superior reanalysis capabilities. The comprehensive coverage offered by GS enables reexamination without the need for additional resampling or resequencing, enhancing diagnostic performance, especially for non-coding regions and areas that are poorly understood.^37,38^ Within this ongoing study, employing adaptive reanalysis of newly identified disease-relevant non-protein-coding regions, such as the *RNU4-2* gene locus, led to the resolution of a previously undiagnosed case.^31,32^

Our results highlight the successful integration of GS into a diagnostic framework traditionally dominated by a series of elaborate methods. The implementation in this prospective study required an extensive training and workflow optimization period for the less experienced genomic teams, addressing challenges posed by the comprehensive data interpretation. Initial discrepancies in expertise between SoC and sGS visible in our learning curve underscored the need for adaptation to the new approaches, better filtering techniques, and platform improvements. Remarkably, for tGS the provided inheritance data helped to offset experience gaps completely and supported more accurate interpretation. Ultimately, through our prospective approach, including a continuous iterative performance comparison, we were able to bridge the experience gap in real-time, refine workflows, and enhance sGS capabilities, thereby achieving diagnostic yields comparable to retrospective analyses.

Of the genetically identified diseases, 15.8% followed autosomal dominant inheritance from a parent, exceeding expectations. This higher rate reflects not only known familial conditions or reduced penetrance but also incomplete parental phenotypic descriptions. In contrast to the anticipated 64% predominance of *de novo* variants, these findings highlight the complexity of inheritance patterns and the need for thorough genetic anamnesis.

The utility of diagnostic solutions depends on both cost and time efficiency. GS’s “one-test-for-all” approach significantly reduces analysis time compared to the sequential SoC methods.^19,39^ A recent study demonstrated that rapid tGS could diagnose rare diseases in 290 critically ill infants in under three days.^39^ In our study, GS facilitated the identification of disease-causing variants, enabling faster clinical decisions and improving diagnostic yield.

However, a direct comparison of our time-to-diagnosis metrics was scientifically infeasible due to differences in study design, analytical strategies, and the varying levels of training and adaptation across the three teams. Notably, tGS eliminated the need for additional segregation analysis, showing a benefit in time even without a formal time-to-diagnosis analysis, making it especially valuable for time-sensitive contexts such as pediatric or acute care, where rapid diagnosis is paramount.

## Conclusion

In summary, our study demonstrated the advantages of integrating short-read GS into rare disease diagnostics, challenging traditional assumptions about diagnostic methods. The unified GS approach improved precision while allowing direct comparisons between tGS and sGS. We demonstrated that tGS offers the highest diagnostic yield, identifying 42.2% of P/LP variants, with notable efficacy in uncovering non-coding, structural, and novel de novo variants. sGS, while slightly less comprehensive, presents a cost-effective alternative, achieving comparable diagnostic success with a diagnostic yield of 41.3%. Both showed potential for further increase in detecting clinically significant variants, matching or surpassing SoC performance.

## Data Availability

All data produced in the present study are available upon reasonable request to the authors

## Supplemental Information

**Supplementary Figure S1.**
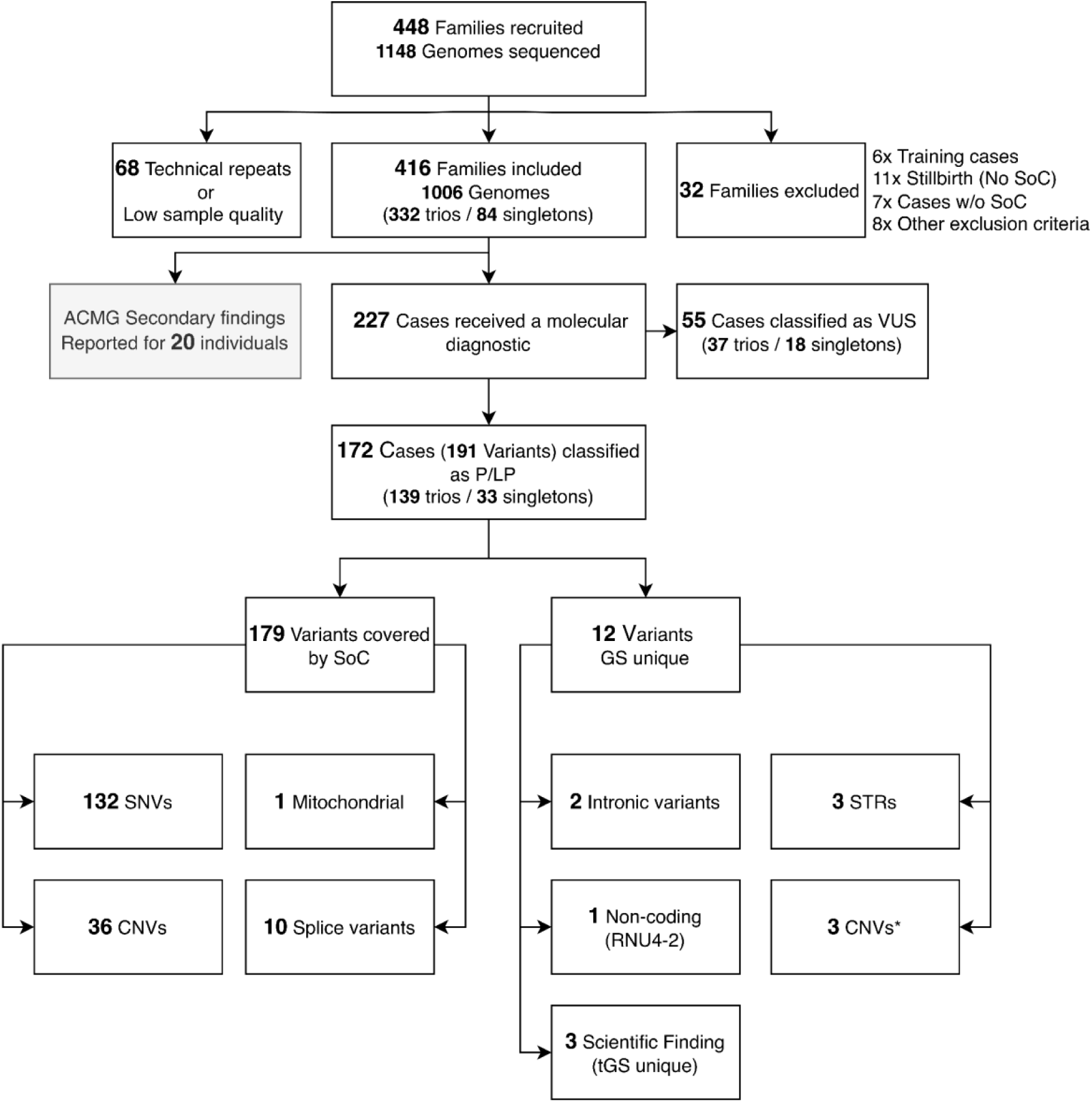
Variant Classification Framework for Cases with a Molecular Diagnosis. Initial recruitment selected 448 cases for sequencing; 32 cases were ultimately excluded. Among the 416 included index cases diagnostic variants were identified in 229 cases. Of these, 173 cases had final diagnostic results with variants classified as pathogenic or likely pathogenic (P/LP), while 56 cases involved variants of uncertain significance (VUS). In total, 191 P/LP variants were identified; 179 were detectable by standard-of-care (SoC) methods, including 36 copy-number variants (CNVs), 132 single-nucleotide variants (SNVs), 10 splicing variants, and 1 mitochondrial variant. An additional 12 variants were identified exclusively through genome sequencing (GS), including 3 intron-to-intron CNVs, 3 short tandem repeats (STRs), 2 deep intronic variants, 1 non-coding variant, and 3 novel *de novo* variants. The latter were identified through GeneMatcher associations made possible by trio genome sequencing (tGS). For 20 individuals we identified an ACMG secondary finding (SF) variant. *Likely detectable by SoC (ES) with improved CNV calling

**Supplementary Figure S2.**
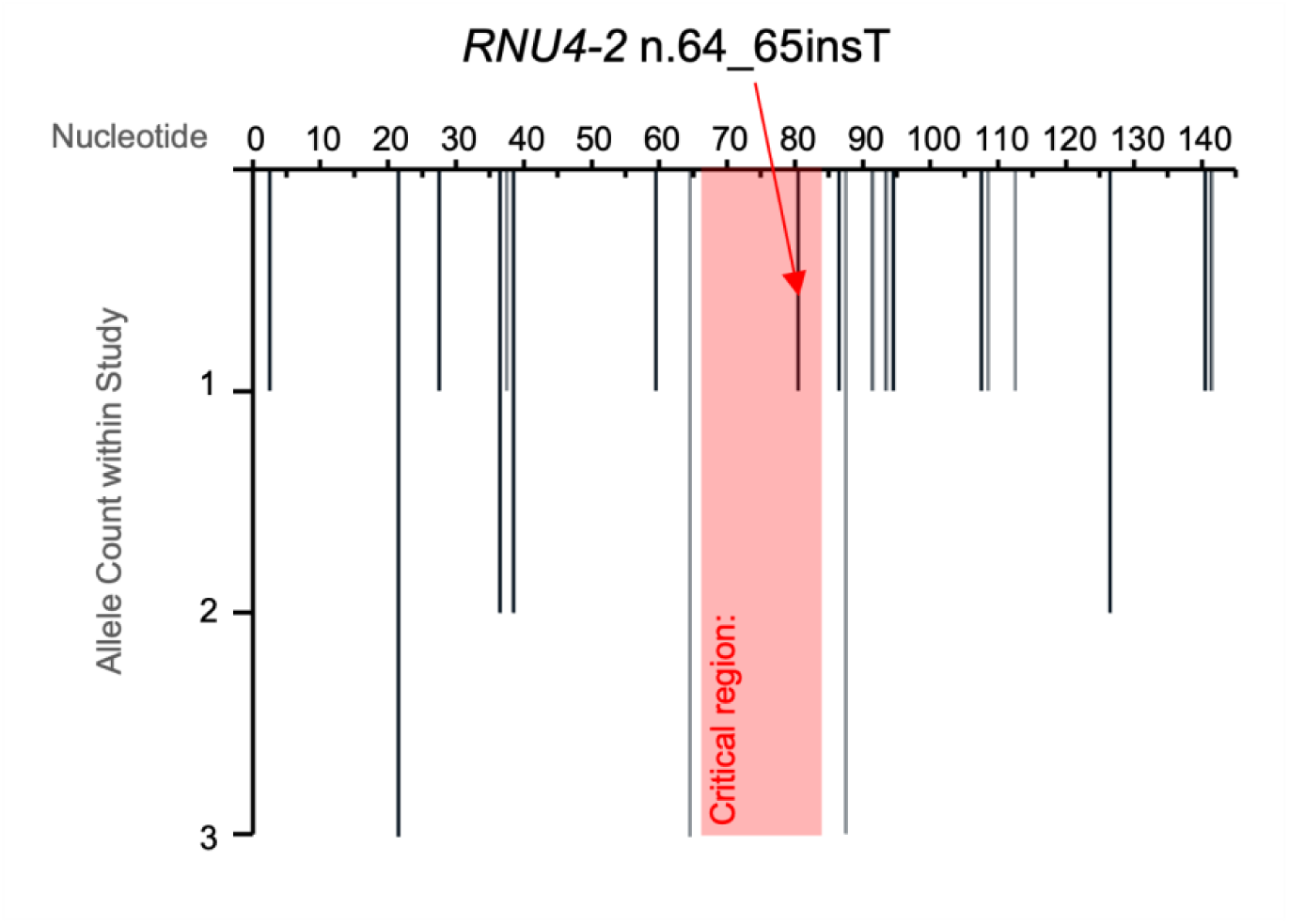
RNU4-2 Reanalysis. The graphic displays allele counts for all variants identified within our cohort in the *RNU4-2* locus, including our reported pathogenic variant, located within the 18 bp critical region.

## Abbreviations

GS: Genome Sequencing
sGS: Singleton Genome Sequencing
tGS: Trio Genome Sequencing
SoC: Standard of Care
array-CGH: Array Comparative Genomic Hybridization
ES: Exome Sequencing
STR: Short Tandem Repeat
CNV: Copy-Number Variant
VUS: Variant of Uncertain Significance
P/LP: Pathogenic / Likely Pathogenic
CADD: Combined Annotation Dependent Depletion
OMIM: Online Mendelian Inheritance in Man
ACMG: American College of Medical Genetics and Genomics
AMP: Association for Molecular Pathology
ACGS: Association for Clinical Genomic Science
SF: Secondary Finding

